# Intravenous Mesenchymal Stem Cells in Extracorporeal Oxygenation Patients with Severe COVID-19 Acute Respiratory Distress Syndrome

**DOI:** 10.1101/2020.10.15.20122523

**Authors:** Sunjay Kaushal, Aisha Khan, Kristopher Deatrick, Derek K. Ng, Abigail Snyder, Aakash Shah, Lina V. Caceres, Ketty Bacallao, Melania Bembea, Allen Everett, Jie Zhu, David Kaczorowski, Ronson Madathil, Ali Tabatabai, Geoffrey Rosenthal, Adriana Brooks, Bangon Longsomboon, Rachana Mishra, Progyaparamita Saha, Yvenie Desire, Russell Saltzman, Kim G.Hankey, Sixto A. Arias, Folusakin Ayoade, Jairo A. Tovar, Rejane Lamazares, Hayley B. Gershengorn, Magali J. Fontaine, Matt Klein, Kristin Mullins, Muthukumar Gunasekaran, Matthias Loebe, Vela Karakeshishyan, Dushyantha T. Jayaweera, Anthony Atala, Ali Ghodsizad, Joshua M. Hare

## Abstract

**Background:** There is an ongoing critical need to improve therapeutic strategies for COVID-19 pneumonia, particularly in the most severely affected patients. Adult mesenchymal stem cell (MSC) infusions have the potential to benefit critically ill patients with acute respiratory syndrome SARS-COV-2 infection, but clinical data supporting efficacy are lacking.

**Methods:** We conducted a case-control study of critically ill patients with laboratory-confirmed COVID-19, severe acute respiratory distress syndrome (ARDS). To evaluate clinical responsiveness in the most critically ill patient we examined outcomes in a sub-group of those requiring extracorporeal membrane oxygenation (ECMO) support. Patients (n=9) were administered with up to 3 infusions of intravenous (IV) MSCs and compared to a local ECMO control group (n=31). The primary outcome was safety, and the secondary outcomes were all-cause mortality (or rate of hospital discharge), cytokine levels, and viral clearance.

**Findings:** MSC infusions (12 patients) were well tolerated and no side effects occurred. Of ECMO patients receiving MSC infusions, 2 out of 9 died (22.2%; 95%CI: 2.8%, 60.0%) compared with a mortality of 15 of 31 (48.4%; 95%CI: 30.2%, 66.9%; p = 0.25) in the ECMO control group. Isolated plasma exosomes containing the SARS-COV-2 Spike protein decreased after MSC infusions between day 14 or 21 after administration (p=0.003 and p=0.005, respectively) and was associated with a decrease in COVID-19 IgG Spike protein titer at same time points (p = 0.006 and p=0.007, respectively). Control ECMO patients receiving convalescent plasma did not clear COVID-19 IgG over the same time frame.

**Interpretation:** Together these findings suggest that MSC IV infusion is well tolerated in patients with a broad range of severity including the most severe COVID-19 ARDS requiring ECMO. These data also raise the possibility that MSCs, in addition to exerting an immunomodulatory effect, contribute to viral clearance and strongly support the conduct of randomized placebo-controlled trial.

## Introduction

The emergence and spread of the novel coronavirus disease 2019 (COVID-2019) has led to an unprecedented global pandemic (1). As of September 11, 2020, COVID-19 has resulted in over 28.5 million cases and ∼900,000 deaths worldwide, and at least 6.4 million cases and ∼190,000 deaths in the United States. The mortality largely results from the development of a severe acute respiratory distress syndrome (ARDS), causing severe fatal hypoxemia, and multisystem organ failure (2). The virus affects the respiratory system by binding to the angiotensin converting enzyme 2 (ACE-2) receptor, which is constitutively expressed in alveolar cells and vascular endothelium(3). Infected patients have a nearly universal development of a severe proinflammatory state, reflected by elevated levels of C-Reactive Protein, Interleukin-6, ferritin, and other cytokines (4-6) (7). Traditional interventions for inflammatory syndromes have had limited efficacy, and there are no therapeutic agents which prevent or treat ARDS in COVID-19 patients, although remdesivir has now entered clinical practice based on studies showing reduced hospitalization days (2, 8, 9). Recently, a Wuhan study reported that out of 201 hospitalized patients, 42% developed ARDS which resulted in a 52% hospital mortality (10). A recent pooled analysis reported as high as 71% mortality in COVID-19 ARDS patients, and a reported 94% mortality in extracorporeal membrane oxygenation (ECMO) COVID-19 patients (11). To reduce mortality in the most severe COVID-19 ARDS patients, mitigating the cytokine storm represents a key strategy. However, many anti-inflammatory drugs have serious side-effects including the potential to offset immune mechanisms aimed at decreasing the viral load. Accordingly, an immunomodulatory therapy, such as the MSC, which also possesses anti-viral properties would be highly valuable in the anti-covid19 armamentarium.

Mesenchymal stem cells (MSCs) possess unique and powerful immunomodulatory characteristics and effectively block cytokine release syndrome in laboratory models(12-14). Recently in China, intravenous adult MSCs have been reported to improve 7 COVID-19 patients with mostly mild symptoms by significantly dampening the cytokine storm and improving pulmonary function and symptoms, without any reported side effects (15). To this point, however, the efficacy of MSC infusion in the most severely ill patients has not been tested, especially in patients requiring extracorporeal membrane oxygenation (ECMO).

Here, we report on an initial experience of patients receiving IV MSC infusions, a subset of whom required ECMO support. Taking advantage of the availability of a local patients with COVID-19 receiving ECMO support (16), we performed a case-control analysis of a cohort of COVID-19 patients receiving intravenous infusions of MSCs following ECMO implantation. Our results extend earlier reports from very sick COVID-19 patients, that MSCs may have salutary effects and are safe.

## Methods

### Study design and patients

This case-control evaluation was part of an overall approach of cell-based therapy at the University of Maryland School of Medicine, Wake Forest University, and University of Miami Health System to treat COVID-19 patients, conducted from April 1, 2020 to September 1, 2020. All patients were confirmed by the real-time reverse transcription polymerase chain reaction (RT-PCR) assay of nasal-pharyngeal swabs for COVID-19. All patients met the inclusion criteria: 1) moderate to severe categories of the Berlin ARDS criteria with a PaO2/FiO2 ≤ 200 mmHg; 2) >18 years of age; exclusion criteria included: 1) nursing mothers; 2) positive bacteria blood cultures; 3) liver transaminases tests ≥5 normal values. Criteria for clinical improvement was determined by weaning off-ECMO support, weaning from mechanical ventilation, or discharge from the hospital. We used single patient emergency-access FDA approval, local Institute Review Board (IRB) approval, and a legal authorized representative for intubated patients provided informed consent.

### Contemporaneous group

Since the MSCs were administered based on compassionate use, there was no randomization. However, we established a control group for comparison of outcomes and clinical characteristics between the MSC-administered subjects and other critically ill ARDS ECMO patients hospitalized at our local institution with similar baseline characteristics. For our institution, we retrospectively characterized all ECMO COVID-19 patients admitted during the same time period from April 1, 2020 to September 1, 2020.

### MSC production

Allogeneic MSCs were derived from healthy bone-marrow donors and the samples were >70% viable at the time of intravenous injection. Patients on ECMO received infusion of allogeneic MSCs via the ECMO circuit return cannula (positioned in the right atrium, with the majority of blood directed across the tricuspid valve and into the pulmonary circulation) and non-ECMO patients received the MSCs intravenously through a central venous catheter. Standard of care infusion was provided for all ARDS patients on or off ECMO.

### Clinical Information

Clinical information for the 12 patients before and after MSC infusion and non-MSC administered control group admitted at the same time was obtained from a review of the hospital electronic medical system and include the following: demographic data, days of admission from symptom onset, and presenting symptoms; data about various infusions, including mechanical ventilation, ECMO support, antiviral therapies, medications, and steroids; clinical data, including PAO2/FiO2, Sequential Organ Failure Assessment (SOFA) score (range 0-24, with higher scores indicating more severe illness), laboratory data, including blood cultures, white blood cell count, chemistry panels assessing liver and kidney function, viral PCR load, inflammatory factors C-reactive protein (CRP; mg/dL), IL-6 (pg/mL), ferritin (ng/mL) and procalcitonin (ng/mL); data from chest imaging studies; and information on complications, such as ARDS, ECMO, MSC infusion reactions, ventilation, bacterial pneumonia, and multiple organ dysfunction syndrome. Since the ECMO blood flow remained constant during support, the oxygen delivery provided by the ECMO circuit remained constant for these patients. Therefore, increasing PAO2/FiO2 ratios or the need to reduce sweep gas flow for these patients is related to improvement in their native lung function.

### Biomarker Analysis

Plasma biomarkers were measured at baseline and post-infusion at selective timepoints. IFN-γ, IL-1β, IL-2, IL-4, IL-6, IL-8, IL-10, IL-12-p70, IL-13, TNF-α, and VEGF-A concentrations(pg/mL) were measured using commercial assay (Meso Scale Discovery, Gaithersburg, MD). To study the imprecision and variability of the biomarker measurements, intra- and inter-assay coefficients of variation (CV) were determined. Intra-assay CVs tested the variability of biomarker measurements performed in the same sample on the same assay plate. Inter-assay CVs tested the variability of biomarker measurements performed in the same sample on different assay plates often used to measure long-term imprecision. When measuring biomarkers with multiplex assays, generally CV’s <15% are targeted and CVs <5% are considered excellent. For the preliminary analysis, all intra-assay CVs were less than 10% and inter-assay CVs for these biomarkers were <10%.

### Detection of SARS-CoV-2 spike antigen containing exosomes

Sera collected from severe COVID-19 patients following MSC IV administration infusion were utilized to detect and quantify SARS-CoV-2 spike antigen containing exosomes by immunoblot. Exosomes were isolated using ultracentrifugation and the presence of exosomes specific marker CD9 was validated using immunoblot as previously published (17-19). Briefly, Total exosome protein (30 μg) was resolved in polyacrylamide gel electrophoresis, and the proteins were transferred into a polyvinylidene difluoride (PVDF) membrane. The PVDF membrane was blocked with 5% non-fat milk prepared in 1x phosphate buffered saline (PBS) and was probed with exosome-specific marker CD9 (312102, BioLegend) and SARS-CoV-2 spike (GTX632604, GeneTex) were used as primary antibodies and goat-anti-mouse conjugated with horse radish peroxidase (7076, Cell Signaling Technology) were used as secondary antibody. The blots were washed with 1x PBS-Tween (Thermo Fisher Scientific), developed using chemiluminescent HRP substrate (WBKLS0500, Millipore Sigma, Burlington, MA), and exposed using Odyssey CLx Imaging System (LI-COR Biosciences, Lincoln, NE). The intensity of SARS-CoV-2 spike antigen was quantified using ImageJ software and normalized with CD9.

### Anti-SARS-CoV-2 IgG ELISA methodology

Anti-SARS-CoV-2 IgG levels were measured in the plasma of COVID 19 patients by following the methodology of Stadlbauer and colleagues (20). 96-well microtiter ELISA plates (Thermo Scientific; Waltham, MA) were pre-coated overnight at 4 ° C with 100 ng/well of the truncated form of spike protein, receptor-binding domain (RBD), dissolved in glycerol and washed (x3) with PBS (Corning; Corning, NY) and 0.05% tween-20 (PBS-T). A 300 uL blocking solution (PBS-T + 5% milk powder (SigmaAldrich, Saint Louis MO) was then added to each well and incubated at room temperature for 1h and washed off before serial dilutions (1:100 to 1:51200) of patients and control plasma samples were added and incubated at room temperature for 1h. The wells were again washed, incubated with 100uL of Anti-Human IgG (Fab specific) HRP-labeled secondary antibody (1:12000) in the dark at 20 ° C for 1h, washed with PBS-T, incubated with 100uL of TMB substrate for 10 minutes followed by 100uL of 1N Sulfuric Acid (SigmaAldrich, Saint Louis MO) to stop the reaction. ELISA plates were then read at absorbance 450nm. Positive signal cutoff was determined using the formula: OD = ((avg OD of negative controls) + 3(SD of negative controls OD)). The negative controls consisted of plasma samples from patients diagnosed with non-SARS-CoV-2.

### Statistical Analysis

To assess temporal changes in cytokines associated with MSC infusions, we constructed graphs anchoring at the baseline cytokine level (on the *x*-axis) which was defined as the sample obtained immediately prior to the first MSC infusion. We plotted cytokine levels 3 days after the first infusion, 3 or 4 days after the second MSC infusion, and 2 days after the third infusion on the *y*-axis. These days were selected based on availability of data and represent reasonably similar durations from MSC infusion to sample collection. All cytokines were plotted in the log scale. To estimate the proportion of samples stable or decreasing after MSC infusions, we created a binary response variable based on the post-MSC sample being less than or equal to the baseline cytokine level and fit a repeated measures logistic model with an independent correlation structure. For each cytokine, we present the proportion of samples less than or equal to baseline levels with corresponding valid 95% confidence intervals. The null hypothesis of this single sample test is that this proportion is equal to 50%. Significantly different estimates at the p= 0.05 level were shown by whether the 95% confidence intervals contained the null. To compare risk of death (i.e., cumulative incidence) in the absence of censoring since all were known to have either died or survived to discharge, we present the proportion who died with exact 95% confidence intervals using the Klopper-Pearson method among those who supported with ECMO for the 9 who received MSC infusions and the 31 who did not receive MSC infusions. Fisher’s exact test were used to compare survival among those who received MSC infusions to those who did not locally, and then in comparison with the ELSO registry. All analyses were conducted in SAS 9.4 software (Cary, NC) and graphs were constructed in R 4.0.0 (Vienna, Austria).

## Results

From April 1,2020 to September 1, 2020, we administered MSCs under FDA eIND to patients (n=12; age range, 39-72 years; 5 women) who met the inclusion criteria (Table1). All MSC administered patients had pre-existing complex medical conditions and one was a former smoker (Supplement Table 1,2). All had received hydroxychloroquine (100%), none had received antivirals and most received corticosteroids (83.33%). Nine patients (75%) were supported with venous-venous ECMO. MSC infusions were administered between 15 and 28 days after hospital admission. During this period, 31 contemporaneous ECMO patients with COVID 19 and similar baseline conditions (table 2) did not receive MSCs and served as a control group (Table 2). No acute-infusion related or allergic reactions were observed within two hours after MSC infusion, including hypotension, increased oxygenation, flushing, or temperature. No delayed hypersensitivity or secondary infections were detected after infusion. Half of the patients received two doses and the other half received three doses every 72 hrs.

**Table 1.**

|  | Pt 1 | Pt 2 | Pt 3 | Pt 4 | Pt 5 | Pt 6 | Pt 7 | Pt 8 | Pt 9 | Pt 10 | Pt 11 | Pt 12 |
| --- | --- | --- | --- | --- | --- | --- | --- | --- | --- | --- | --- | --- |
| Age Range (years) | Ranged 39-72 years |  |  |  |  |  |  |  |  |  |  |  |
| Gender | 5 Females, 7 Males |  |  |  |  |  |  |  |  |  |  |  |
| Weight (kg) | 60 | 140 | 62 | 84.2 | 94.8 | 155 | 100 | 70.5 | 105 | 98 | 239.04 | 80 |
| Smoker? | No | No | No | Former | No | No | No | No | Yes | No | Unknown | No |
| Symptoms | Fever; Cough; Dyspnea; Nausea/Emesis; Pharyngitis; Headache | Fever; Cough; Dyspnea | Fever; Cough; Dyspnea; Nausea/Emesis; Severe Chest Pain; Abdominal Pain; Myalgias; Tachypnea | Fever; Cough; Dyspnea | Fever; Cough; Myalgias; Dyspnea | Cough; Dyspnea | Fever; Cough; Malaise; Dyspnea | Fever; Headache; Malaise; Nausea/Emesis; Diarrhea; Dyspnea; Cough | Fever; Dyspnea | Fever; Myalgias; Dyspnea | Dyspnea | Dyspnea |
| COVID-19 Exposure | Travel | Community acquired | Community acquired | Community acquired | Community acquired | Unknown | Unknown | Community acquired | Unknown | Community acquired | Unknown | Unknown |
| Estimated Incubation Period (days) | 4-5 days | No Known Exposure | 3 days | No Known Exposure | No Known Exposure | No Known Exposure | No Known Exposure | No Known Exposure | No Known Exposure | No Known Exposure | No Known Exposure | No Known Exposure |
| Time from Symptom Onset to Hospital Admission (days) | 4 | 10 | 7 | 8 | 5 | 2 | 7 | 14 | 6 | 7 | 3 | 3 |
| Time from Admission to First MSC Infusion (days) | 17 | 15 | 22 | 20 | 21 | 28 | 41 | 16 | 16 | 21 | 7 | 5 |
| Complications Prior to MSC Infusion | Pneumonia; Hypoxic Respiratory Failure; Hypotension; Right Ventricular Strain; Pulmonary Hypertension | Pneumonia; Bradycardic/hypoxic arrest; Right Ventricular Failure; SVT; Hypotension; Pneumothorax | Pneumonia; Refractory Hypoxic and Hypercarbic Respiratory Failure; Delirium; Hypotension; Pneumothorax; Leukocytosis; Thrombocytosis; Coagulopathy | Pneumonia; Hypoxic Respiratory Failure; Acute Kidney Injury; ST Elevation Myocardial Infarction; Brief VT Arrest; Ileus | Pneumonia; Hypoxemic Respiratory Failure | Pneumonia; Hypoxemic Respiratory Failure | Pneumonia; Acute Hypoxic Respiratory Failure | Pneumonia; Hypoxic Respiratory Failure; Hypotension; Delirium; Anemia; Hematuria | Pneumonia; Hypoxic Respiratory Failure; Altered mental status; Tachycardia; Sepsis; Bacterial pneumonia (strep); Hyperglycemia Adrenocortical insufficiency; Anemia; Thrombocytopenia; Coagulopathy | Pneumonia; Acute hypoxemic respiratory failure; Rapid Atrial fibrillation; Heparin-induced thrombocytopenia | Possible Community Acquired Pneumonia; Hypoxic Respiratory Failure; ST-Elevation Myocardial Infarction | Pneumonia; Hypoxic Respiratory Failure |
| Oxygen Therapy at Infusion | Mechanical Ventilation; VV ECMO | Mechanical Ventilation; VV ECMO | Mechanical Ventilation; VV ECMO | Mechanical Ventilation | Mechanical Ventilation | Mechanical Ventilation; VV ECMO | High Flow Nasal Cannula | Mechanical Ventilation; VV ECMO | Mechanical Ventilation; VV ECMO | Mechanical Ventilation; VV ECMO | Mechanical Ventilation; VV ECMO | Mechanical Ventilation; VV ECMO |
| Disease Class | Critical | Critical | Critical | Critical | Critical | Critical | Critical | Critical | Critical | Critical | Critical | Critical |
| Infusion | Hydroxychloroquine | Hydroxychloroquine | Hydroxychloroquine; Tocilizumab | Hydroxychloroquine | Hydroxychloroquine; Tocilizumab | Hydroxychloroquine; Tocilizumab | Hydroxychloroquine; Tocilizumab | Convalescent Plasma | Acalabrutinib; Remdesivir | Convalescent Plasma; Remdesivir | Hydroxychloroquine | Convalescent Plasma; Tocilizumab |
| Steroid | None | Methylprednisolone | Methylprednisolone | Methylprednisolone | Hydrocortisone; Dexamethasone; Prednisolone | Dexamethasone | Methylprednisolone | Methylprednisolone | Methylprednisolone | Methylprednisolone | NA | N/A |
| Number of MSC Infusions | 3 | 3 | 3 | 2 | 2 | 2 | 3 | 3 | 3 | 3 | 3 | 3 |

**Table 2.**
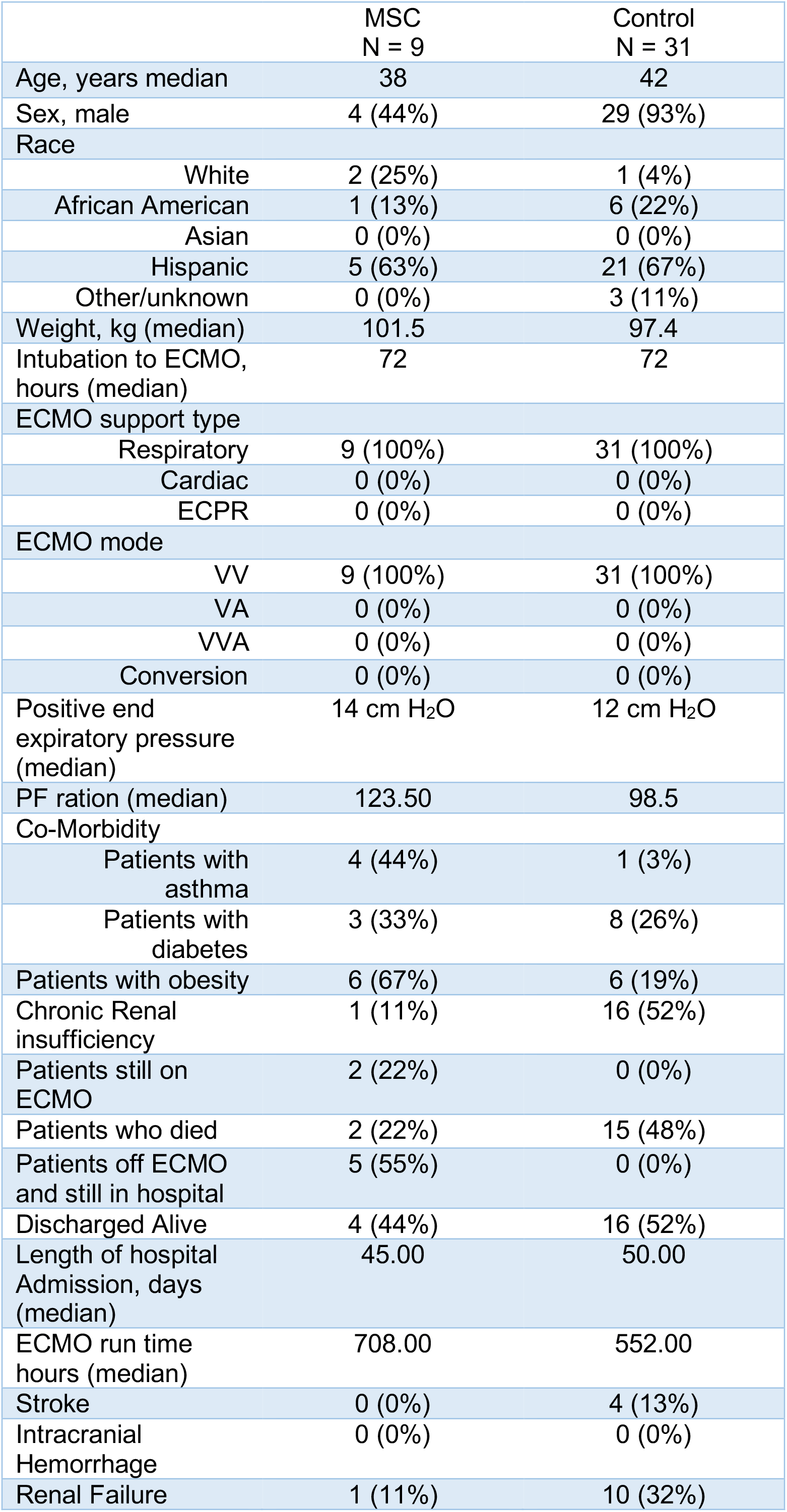
Clinical Characteristics of COVID-19 Patients Who Received MSC infusion.

|  | MSC<br>N = 9 | Control<br>N = 31 |
| --- | --- | --- |
| Age, years median | 38 | 42 |
| Sex, male | 4 (44%) | 29 (93%) |
| Race |  |  |
| White | 2 (25%) | 1 (4%) |
| African American | 1 (13%) | 6 (22%) |
| Asian | 0 (0%) | 0 (0%) |
| Hispanic | 5 (63%) | 21 (67%) |
| Other/unknown | 0 (0%) | 3 (11%) |
| Weight, kg (median) | 101.5 | 97.4 |
| Intubation to ECMO,<br>hours (median) | 72 | 72 |
| ECMO support type |  |  |
| Respiratory | 9 (100%) | 31 (100%) |
| Cardiac | 0 (0%) | 0 (0%) |
| ECPR | 0 (0%) | 0 (0%) |
| ECMO mode |  |  |
| VV | 9 (100%) | 31 (100%) |
| VA | 0 (0%) | 0 (0%) |
| VVA | 0 (0%) | 0 (0%) |
| Conversion | 0 (0%) | 0 (0%) |
| Positive end<br>expiratory pressure<br>(median) | 14 cm H<sub>2</sub>O | 12 cm H<sub>2</sub>O |
| PF ration (median) | 123.50 | 98.5 |
| Co-Morbidity |  |  |
| Patients with<br>asthma | 4 (44%) | 1 (3%) |
| Patients with<br>diabetes | 3 (33%) | 8 (26%) |
| Patients with obesity | 6 (67%) | 6 (19%) |
| Chronic Renal<br>insufficiency | 1 (11%) | 16 (52%) |
| Patients still on<br>ECMO | 2 (22%) | 0 (0%) |
| Patients who died | 2 (22%) | 15 (48%) |
| Patients off ECMO<br>and still in hospital | 5 (55%) | 0 (0%) |
| Discharged Alive | 4 (44%) | 16 (52%) |
| Length of hospital<br>Admission, days<br>(median) | 45.00 | 50.00 |
| ECMO run time<br>hours (median) | 708.00 | 552.00 |
| Stroke | 0 (0%) | 4 (13%) |
| Intracranial<br>Hemorrhage | 0 (0%) | 0 (0%) |
| Renal Failure | 1 (11%) | 10 (32%) |

Follow-up data for all outcomes were available through September 1, 2020. No patients were lost to follow-up for the primary outcome of safety with MSC infusion which showed no reported side effects. To assess whether MSC-therapy could have a clinical benefit, we compared the mortality rate of infused patients to a relevant contemporaneous group. Of ECMO patients who did not receive MSC infusion, 15 of 31 (48.4%; 95%CI: 30.2%, 66.9%; p=0.25) died compared with a mortality rate of 2 of 9 patients receiving MSC infusion died (22.2%; 95%CI: 2.8%, 60.0%).

Six patients were successfully decannulated from ECMO on post-MSC infusion day 4, 15, 16, 18, 26, and 28 days, respectively, and of these, 2 patients required a tracheostomy and 5 patients were discharged home. The two tracheostomy patients had their tracheostomy decannulated. One patient remained on ECMO at the end of the follow-up period, and two patients died on ECMO after MSC infusion.

Two of 3 MSC administered patients not on ECMO were extubated and discharged from the hospital with length of stay 34 and 67 days. A third MSC administered patient not on ECMO suffered a myocardial infarction that resulted in a left ventricle ejection fraction of 10%, cardiogenic shock and died on hospital day 29 which was 9 days after MSC infusion.

The SOFA score ranged from 4 to 19 prior to MSC infusion, and decreased to a range of 2 to 15 at 14 days following infusion (Supplement Table 1 and Figure 1). The PaO2/FiO2 ranged from 78.33 to 371.43 prior to MSC infusion, and increased for most patients (83.33%), ranging from 109 to 503.57 14 days after MSC infusion (Supplement Table 1 and Figure 1). After infusion, inflammatory biomarkers concentrations for CRP, IL-6, ferritin, and procalcitonin decreased in all patients (Supplement Table1 and Figure 2). Patient chest radiographs all demonstrated severe bilateral dense pulmonary parenchymal opacities prior to MSC infusion and showed improvement of the pulmonary opacities on the third day after MSC infusion and gradual resolution 10 days after the first MSC dose (Supplement Figure 3). Even Patient 2 and Patient 4, who eventually died, had resolving chest radiograph findings (Supplement Figure 3).

**Figure 1.**
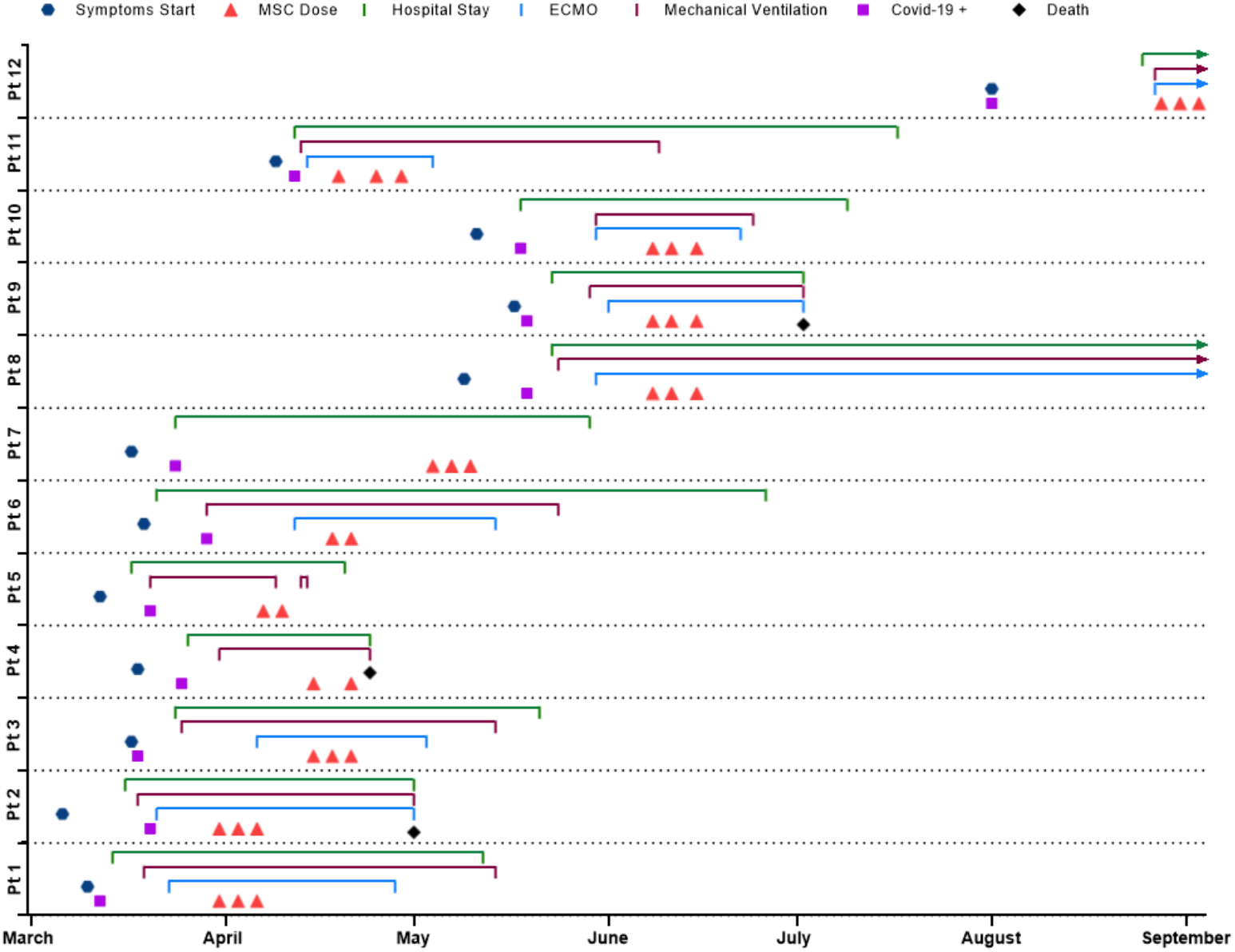
Schematic description of 12 cases of COVID-19. COVID-19= coronavirus disease 2019. ECMO=extracorporeal membrane oxygenation.

**Figure 2.**
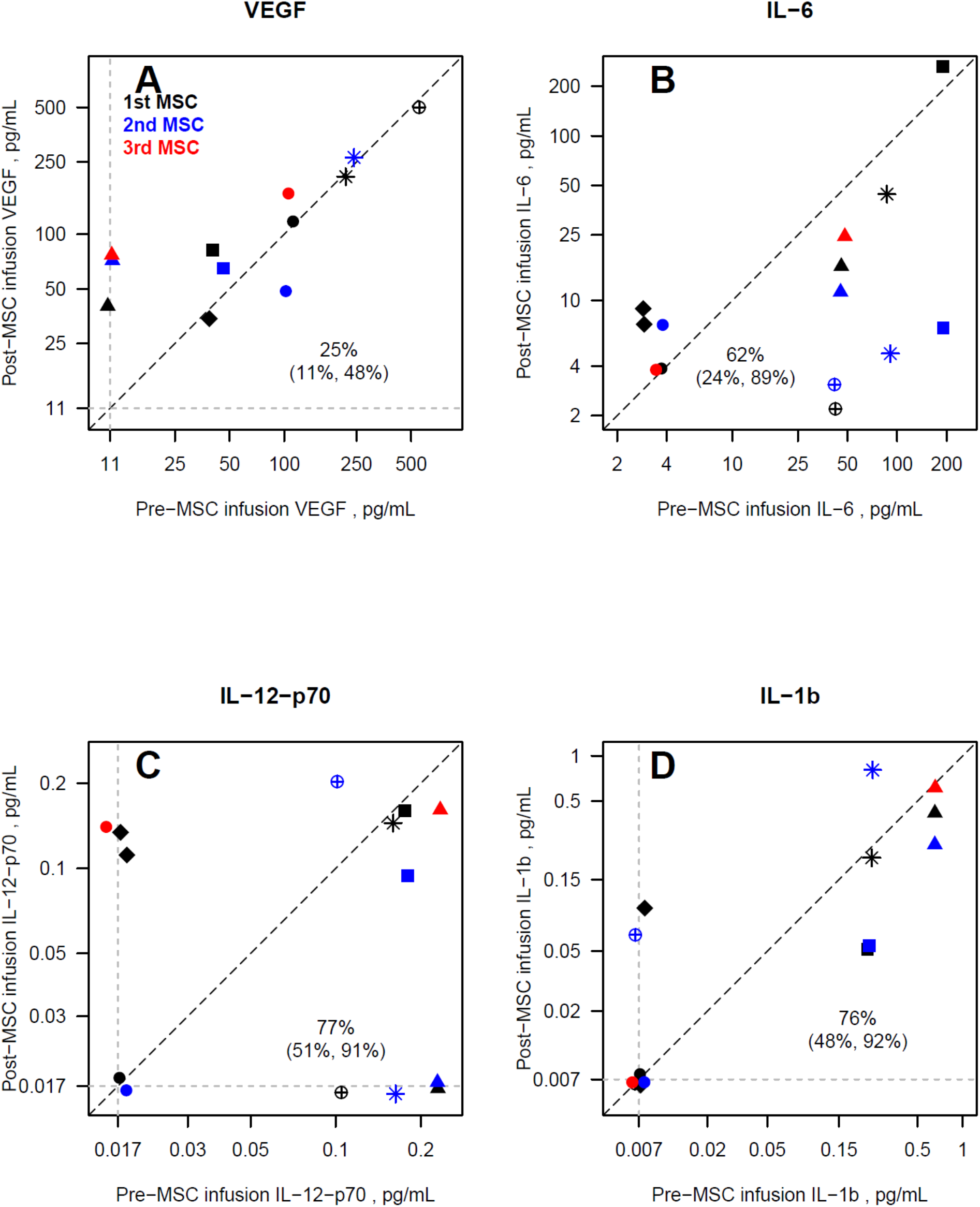
Changes in multiplex biomarkers relative to baseline defined as concentration of the blood sample obtained immediately prior to MSC infusion (x-axis) and subsequent collections (y-axis) for VEGF-A (5A), interleukin-6 (IL-6; 5B), Interleukin-12-p70 (IL-12-p70; 5C) and Interleukin-1b (IL-1b; 5D) in pg/mL. Point shapes represent individuals receiving the MSC infusion (n= 6). Black points represent samples 3 days after the first MSC infusion (n= 6), blue points represent samples taken 3 or 4 days after the second MSC infusion (n= 5), red points represent samples taken 2 days after the third MSC infusion (n= 2). The dark dash line of identity indicates no change from baseline. Dashed grey lines represent lower limits of detection for each biomarker and all data were within the limits of detection. Proportions (with 95% confidence intervals) of post-MSC infusion biomarker that were less than or equal to baseline biomarker levels are presented based on logistic regression models using generalized estimating equations with an independent correlation structure to account for within-person repeated measures.

### Inflammatory Cytokines

We next examined the levels of 11 pro-inflammatory and anti-inflammatory cytokines from plasma samples taken at baseline and approximately three days after each MSC infusion by examining their ratio of change from baseline and after each MSC infusion (Figure 2, Supplement Figure4). These samples were grouped as 3 days after first infusion (black; n= 6), 3 or 4 days after the second MSC infusion (blue, n= 5), and 2 days after the third infusion (red, n= 2). VEGF-A in 25% of post-MSC infusion samples was less than or equal to baseline levels (95% CI: 11%, 48%) indicating that VEGF-A was increased in 75% of post-infusion. This was significantly different from the null of 50% (p= 0.035). For the remaining cytokines, 61% to 77% of samples decreased, apart from IL-8 in which 46% of the samples decreased. These were not significantly different from the null, except for IL-12-p70 in which 77% were decreased (95% CI: 51%, 91%; p= 0.043).

### COVID-19 spike protein and antibody titers

Immunoblot analysis showed that the COVID 19 spike protein antigens were present at baseline in isolated plasma exosomes from COVID 19 patients prior to MSC infusion and significantly reduced at post-infusion day 14 (p=0.007) in 5 patients or at day 21 (p=.006) in 3 patients. Of the three patients, specificity of the exosome assay was demonstrated in three negative COVID 19 patients which revealed absence of spike protein in isolated plasma exosomes. SARS-Co V-2 antibody levels measured in plasma samples drawn pre- and post-MSC infusion in COVID 19 patients (Figure 4). SARS-CoV-2 antibody against a truncated spike protein RBD was detected in all patients at time baseline prior to MSC infusion and its titer was significantly decreased at day 14 post infusion, unchanged there until day 21. In comparison, ECMO patients receiving convalescent plasma did not reveal similar reductions over the 14 to 21 day period (data not shown).

**Figure 3.**
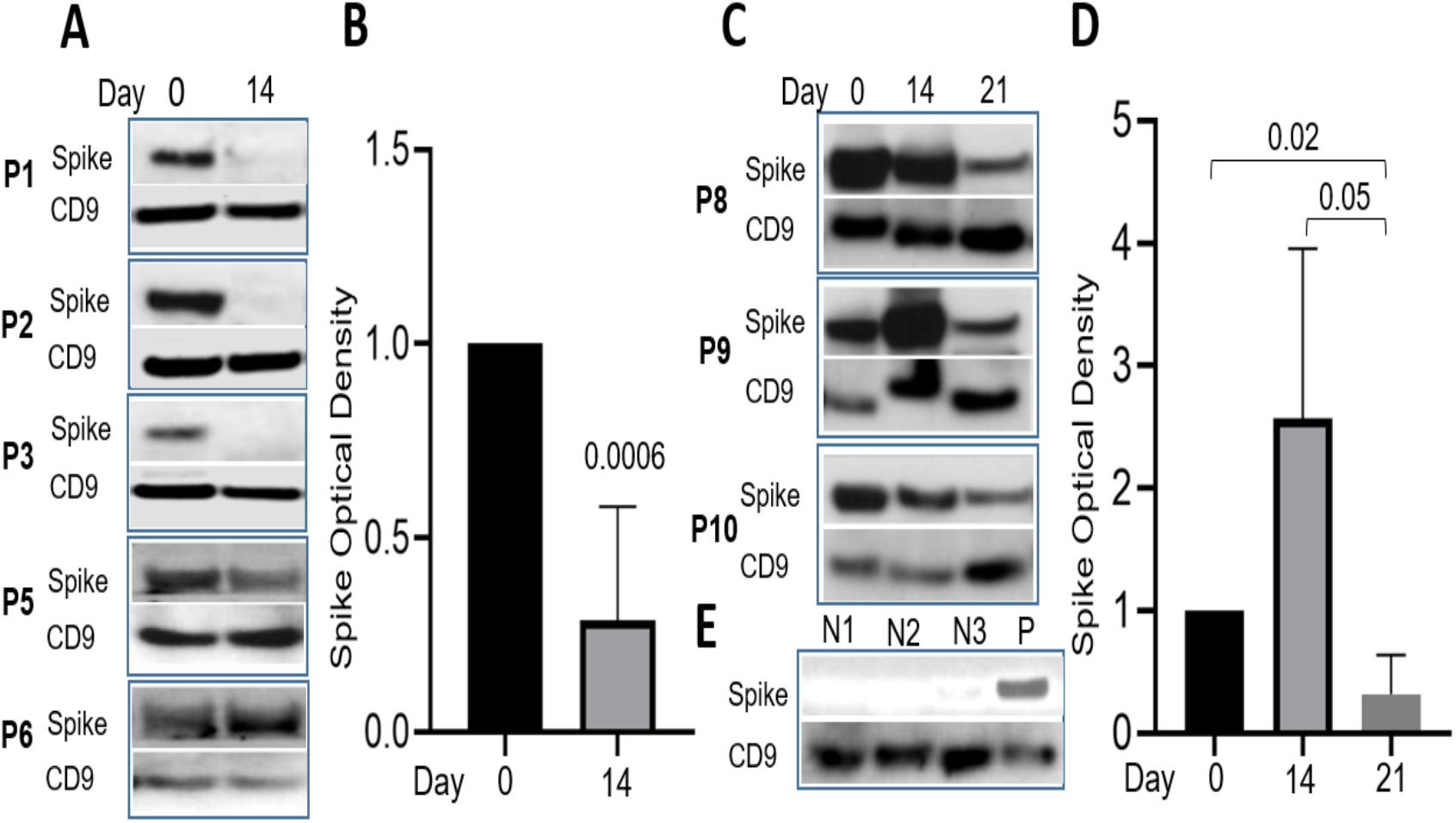
SARS-CoV-2 spike antigen was demonstrable in exosomes isolated from sera of COVID-19 patients: **A)** Immunoblot results showed the presence of SARS-CoV-2 spike antigen on sera collected on day 0 (pre-treatment) and significantly reduced on day 14 (post-treatment) following MSCs therapeutic intervention. **B)** The intensity of SARS-CoV-2 spike antigen and exosome specific marker CD9 was quantified using image J and histogram represents significant (p=0.006) reduction of SARS-CoV-2 spike antigen level on day 14 following MSCs treatment. **C)** Sera collected from additional COVID-19 patients treated with MSCs on day 0 and 14 showed the presence of SARS-CoV-2 spike antigen and significant reduction (p=0.05) compared on day 21. **D)** Graphical representation shows significant reduction of SARS-CoV-2 spike antigen on day 21 compared to sera collected on day 0 and 14 from COVID-19 patients. **E)** Immunoblot results shows the absence of SARS-CoV-2 spike antigen in exosomes collected from healthy subjects negative (N1 & N2) for COVID-19 and COVID-19 patient (P) derived exosomes served as positive control. Optical density of SARS-CoV-2 spike antigen and exosomes specific marker CD9 were measured using ImageJ software (NIH). The level of SARS-CoV-2 spike antigen in the exosomes isolated from COVID-19 patients with MSCs treatment was compared among day 0, 14 and 21 using non-parametric, two tailed T-test. Spike, SARS-CoV-2 spike antigen, P1 to P10, COVID-19 patients; N1, N2 & N3, COVID-19 negative healthy subjects; P, COVID-19 positive patient.

**Figure 4.**
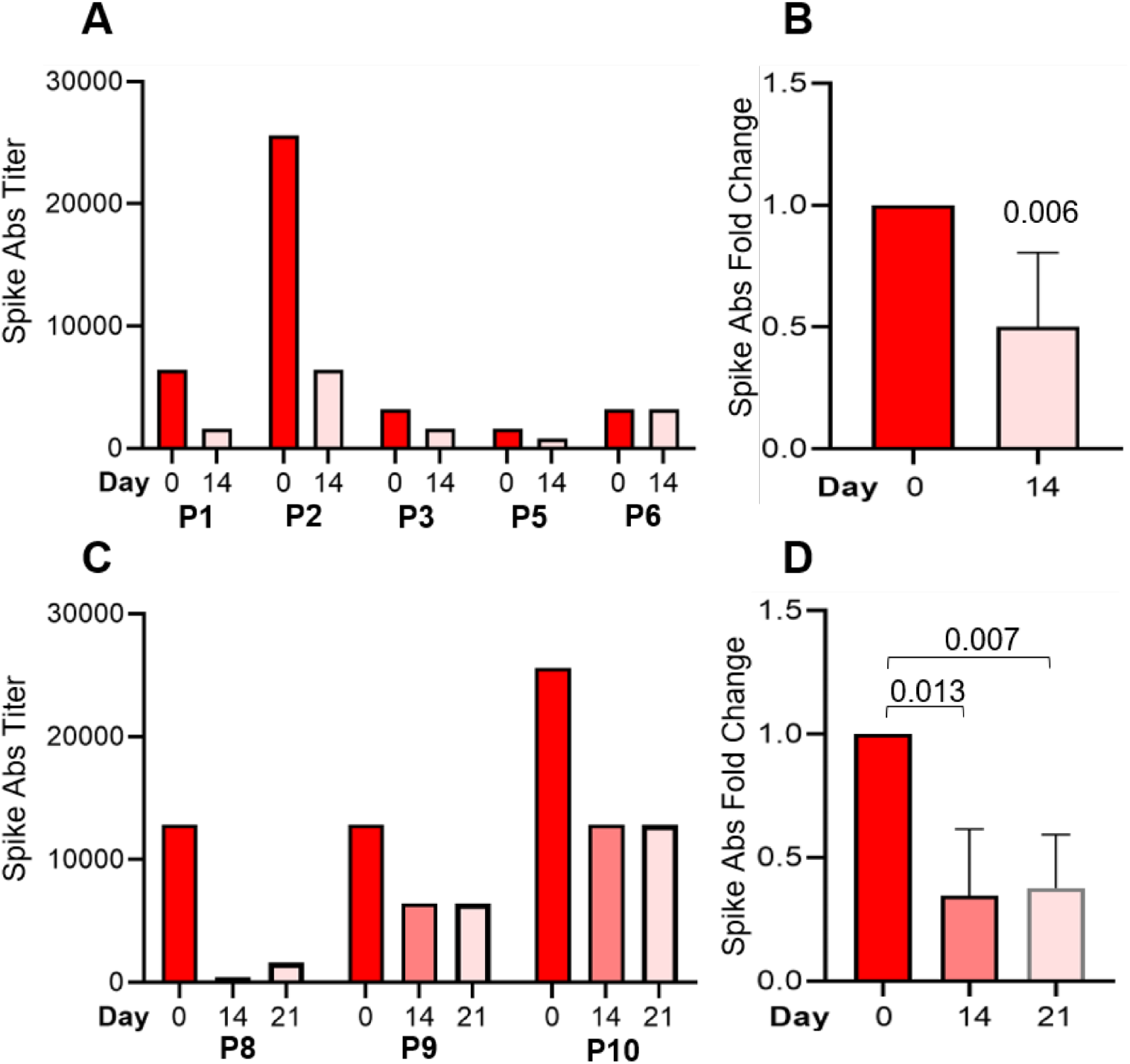
SARS-Co V-2 antibody titer and fold change in plasma pre- and post-MSC infusion on days 0 and 14 respectively in patients P1-P6 (n=6) (A,B); and in plasma pre- and post-MSC infusion on days 0, 14, and 21 in patients P7-P10 (n=3). Fold change was evaluated using student t test and considered significant with p<0.05.

## Discussion

In this case-control study, 12 critically ill COVID-19 patients were administered intravenous infusions of culture-expanded allogeneic MSCs. In response, inflammatory cytokines declined within days of MSC infusion, patients improved clinical status, SOFA scores, and PaO2/FiO2 ratios, and exhibited resolution of COVID-19 pneumonia on chest radiographs. Among the sickest cohort requiring ECMO support, mortality was numerically less than that of the global world-wide experienced as tabulated by the registry. Of 7 surviving ECMO patients, 6 have been successfully decannulated with one subject remaining on ECMO at study endpoint. These findings, which are preliminary, nevertheless add to other early stage reports, substantiate the safety of these infusions, and strongly support efforts to rigorously test this strategy in placebo-controlled trials.

The novel SARS-CoV2 virus can induce ARDS as a serious manifestation of COVID-19 which can rapidly progress to refractory pulmonary failure. In the most advanced cases, ECMO support may be considered as a rescue therapy, with limited prognosis for viral infections in general. For instance, ECMO support for ARDS in patients with Middle East Respiratory Syndrome Coronavirus (MERS-CoV) reduced the in-hospital mortality rate to 65% and decreased length of intensive care unit stay when compared to conventional therapy(16). However, a pooled analysis of early ECMO support in COVID-19 patients reported a 94% in-hospital mortality rate as compared to 70.9% with conventional therapy (11). Query of the Extracorporeal Life Support Organization (ELSO) registry as September 15, 2020, demonstrated that 895 COVID-19 patients have required ECMO support with the majority still on ECMO, and 51% in-hospital mortality (www.elso.org). In our study, we report survival and clinical improvement in 10 out of 12 COVID-19 patients who were administered with MSCs.

Despite encouraging efficacy of MSC administration in pre-clinical models of viral infection, there are limited published clinical data available for various viral infections. For instance, MSC infusion in ARDS induced by epidemic Influenza A (H7N9) had a reduced mortality to 17.6% when compared to non-administered control with a mortality of 54.4% (17). More specifically for COVID-19, a Chinese study of 7 patients reported that the intravenous administration of adult MSCs improved the clinical status and oxygenation, reduced inflammation, increased anti-inflammatory cytokines, and led to the discharge of all administered patients(15). Even so, the major limitation of the latter study is that the 7 COVID-19 patients were not categorized as having severe ARDS, rather they had at most mild ARDS.

In our study, we administered MSCs to 12 patients admitted to hospital with severe ARDS, 9 of whom had a requirement for ECMO support. MSC intravenous administration had no serious adverse effects. Timing of MSC administration may be critical to achieve a more robust clinical response as most of the patients received MSCs late in their hospital course. This report offers an important clinical insight as all ongoing or planned clinical trials for cell-based infusions excludes ECMO COVID-19 patients who are the most critically ill and therefore challenges the therapeutic MSC intervention to its highest capacity. Considering the high mortality of COVID-19 ECMO administered patients, MSC infusion needs to be further evaluated in this highly selective gravely ill patient population as well as less severe ARDS patients.

This study has important limitations that warrant mention. First, this was a small clinical experience of 12 patients that lacked a randomized control group. Nonetheless the ability to compare mortality with a contemporaneous control group and a world-wide global experience is supportive of the potential clinical merits of this approach. In addition, the change in PaO_2_/FiO_2_, SOFA and VEGF-A and IL12p70 levels after MSC infusion provided encouraging positive endpoints that warrant further investigation of MSC infusion. Second, all patients received additional therapies during their hospital stay which may have influenced patient outcomes independently of or additive to the MSC infusion. Third, this case series cannot discern whether this infusion will reduce in-hospital mortality if scaled up for broad clinical use, although this analysis provides an intriguing summary patient response in North America that are fairly consistent with reports from China.

In this preliminary series of 12 critically ill patients with COVID-19 pneumonia, most requiring ECMO support, clinical status improved after MSC infusion. While this study only reports results from relatively few patients, this experience suggests that a patient population with great unmet need, like ECMO patients, who are often excluded from current clinical trial design should be studied in randomized, placebo-controlled trials. MSCs may represent a potentially safe anti-inflammatory therapy that can contribute to patient recovery in the most severe forms of COVID-19 pneumonia.

## Data Availability

All data will be provided upon written request

## Author Contributions

Drs. Kaushal and Hare had full access to all of the data in the study and take responsibility for the integrity of the data and the accuracy of the data analysis.

As the Director of ISCI (one of the sponsors) and the co-leader of this project, Dr. Hare had full access to the study design, interpretation of the data, preparation, review and approval of the manuscript, and decision to submit the manuscript.

As the Director of Pediatric Cardiac Surgery at the University of Maryland (one of the sponsors) and the co-leader of this project, Dr. Kaushal had full access to the study design, interpretation of the data, preparation, review and approval of the manuscript, and decision to submit the manuscript.

*Concept and design:* Kaushal, Khan, Deatrick, Rosenthal, Dushyantha, and Hare

*Acquisition, analysis, or interpretation of data:* Kaushal, Deatrick, Ng, Snyder, Shah, Caceres, Bembiea, Everett, Zhu, Saltzman, Tovar, Karakeshishyan, and Hare

*Drafting of the manuscript:* Kaushal, Hare, Ng, and Snyder

*Critical revision of the manuscript for important intellectual content:* Kaushal, Khan, Deatrick, Ng, Snyder, Bembiea, Everett, Tabatabai, Hankey, Arias, and Hare

*Statistical analysis:* Kaushal, Ng, Snyder, and Bembiea

*Obtained funding:* Kaushal and Hare

*Treating physicians:* Kaushal, Deatrick, Kaczorowski, Madathil, Tabatabai, Ghodsizad, Arias, Ayoade, Gershengorn, and Loebe

*Administrative, technical, or material support:* Khan, Snyder, Shah, Caceres, Bacallao, Brooks, Longsomboon, Mishra, Saha, Desire, Saltzman, Hankey, Tovar, Lamazares, and Karakeshishyan

## Role of Funder/Sponsor

The funding agencies had no role in the design and conduct of the study, interpretation of the data, preparation, review, or approval of manuscript and decision to submit the manuscript.

Information about Patients:

- Intubation to ECMO hours, Positive end expiratory pressure, stroke, renal failure, renal insufficiency, intracranial hemorrhage, and intubation to ECMO hours was not included for one patient with MSC infusion from Miami
- Any patient that was remained in the hospital were not accounted for in the length of hospital admission.
- Two patients remained on ECMO and was not accommodated in ECMO run time hours
- ECMO run time was calculated by the number of days on ECMO times 24 hours

## Supplementary Data

**Supplemental Table 1.**

|  | Patient 1 | Patient 2 | Patient 3 | Patient 4 | Patient 5 | Patient 6 | Patient 7 | Patient 8 | Patient 9 | Patient 10 | Patient 11 | Patient 12 |
| --- | --- | --- | --- | --- | --- | --- | --- | --- | --- | --- | --- | --- |
| SOFA Score<sup>d</sup> |  |  |  |  |  |  |  |  |  |  |  |  |
| Baseline | 9 | 14 | 4 | 10 | 4 | 19 | NA | 5 | 13 | 6 | NA | NA |
| Day 1 | 8 | 16 | 4 | 17 | 6 | 20 | 2 | 6 | 10 | 9 | NA | 8 |
| Day 3 | 9 | 19 | 3 | 13 | 3 | 17 | 2 | 5 | 12 | 7 | NA | 9 |
| Day 5 | 11 | 19 | 3 | 13 | 3 | 18 | NA | 5 | 10 | 8 | NA | 9 |
| Day 7 | 11 | 18 | 3 | 13 | 2 | 18 | 2 | 5 | 6 | 11 | NA | 10 |
| Day 14 | 2 | 10 | 2 | NA | 2 | 15 | NA | NA | NA | NA | NA | 10 |
| PaO<sub>2</sub>:FiO<sub>2</sub> Ratio<sup>e</sup> |  |  |  |  |  |  |  |  |  |  |  |  |
| Pre-Intubation | 57.16 | 65 | 120 | 220.4 | 192.5 | 46 | NA | NA | 116.4 | 70 | NA | 198 |
| Day 0 | 224 | 78.33 | 371.43 | 240 | 141.38 | 125.56 | 30.77 | 160 | 65.00 | 236.67 | NA | 148 |
| Day 1 | 230 | 122 | 376.19 | 114 | 298.00 | 148.89 | 188.89 | 188.3 | 87.00 | 263.33 | NA | NA |
| Day 3 | 225 | 104.29 | 519.05 | 178 | 245.25 | 104 | 280.55 | 237.5 | 188 | 197.5 | NA | 9 |
| Day 5 | 370 | 96.25 | 419.05 | 207.5 | 157.25 | 125 | NA | 217.5 | 145 | 240 | NA | 9 |
| Day 7 | 386.67 | 145 | 419.05 | 182.5 | 277.60 | 95.71 | NA | 386.67 | 218 | 266.67 | NA | 10 |
| Day 14 | 407.5 | 210 | 503.57 | NA | 269.375 | 109 | NA | 177.5 | 93 | 185 | NA | 10 |
| Time from Confirmed COVID Positive Result to First MSC Treatment (days) | 19 | 11 | 28 | 21 | 18 | 20 | 41 | 20 | 20 | 21 | 7 | 57 |
| First Negative COVID Result Post MSC Treatment (days) | 15 | 6<sup>a</sup> | 23 | No negative test | 1/8<sup>b</sup> | 26/32 | 6 | No negative test to date | No negative test to date | No negative test to date | 47 | 8 |
| Mechanical Ventilation/ECMO Days Prior to MSC Treatment | 12/8 | 13/10 | 21/9 | 15/NA | 18/NA | 20/6 | N/A | 15/9 | 10/7 | 9/9 | 6/5 | 39/29 |
| Current Oxygen Therapy | Room Air | NA | Room Air | NA | Room Air | Room Air | Room Air | Mechanical Ventilation; ECMO | N/A | Mechanical Ventilation | Mechanical Ventilation | Mechanical Ventilation ECMO |
| Decannulation Day Post MSC Treatment | 28 | NA | 18 | NA | N/A | 26 | NA | NA | NA | 4 | 15 | NA |
| C-Reactive Protein (mg/dL) |  |  |  |  |  |  |  |  |  |  |  |  |
| Baseline | 6.6 | 39.5 | 0.7 | 1.8 | 16.1 | 37.4 | 1 | 19.1 | 36.3 | 13.7 | 114.3 | 3.3 |
| Day 3 | 4 | 25.6 | 0.5 | 8.9 | 10.2 | 36.1 | 0.9 | 14.8 | 37.7 | 6.3 | 65.7 | 2.4 |
| Day 6 | 3.8 | 8.2 | 6.6 | 3.4 | 6.5 | 28 | 0.6 | 7.4 | 31.1 | 9 | 196.5 | 0.9 |
| Day 14 | 2.9 | 3.8 | 5.1 | NA | NA | 26.1 | 1.1 | 6 | 7.7 | 33.1 | 268.5 | 1.1 |
| Interleukin-6 (pg/mL) |  |  |  |  |  |  |  |  |  |  |  |  |
| Baseline | 7 | 33 | 146 | 5 | 495.5 | 100 | 5.28 | 33.2 | 39.8 | <2 | 9.2 | 14.3 |
| Day 3 | 5 | 15 | 46 | 9 | 19.7 | 71.59 | 4.9 | 7.4 | 16.2 | 6 | 8.5 | 12.09 |
| Day 6 | 5 | 5 | <5 | <5 | 256.7 | 29.42 | 13.53 | 46.7 | 6.4 | 7 | 26.1 | NA |
| Day 14 | <5 | <5 | 6 | NA | 1.9 | 67.4 | 4.57 | 52 | 42.3 | 34.8 | NA | NA |
| Length of Hospital Stay (days) | 59 | 46 | 58 | 29 | 34 | 96 | 67 | Still admitted; Day 93 | 33 | 41 | 74 | Still admitted Day 64 |
| Current Status as of June 25, 2020 | Discharged Home | Deceased | Discharged Home | Deceased | Discharged Home | Stable; On floor | Discharged Home | ICU, Remains on ECMO Support | Deceased | Discharged Home | Discharged Home | ICU, Remains on ECMO support |

a; Retested positive day 7, no other testing done to date. b; Retested positive day 5, followed by negative results on days 8 and 11. c; Reintubated day 6, extubated day 7. Values are +/- 1 day, except for day 14 values which are +/- 3 days d. Retested positive day 29, followed by negative results on day 32 to current

**Supplemental : Table 2.**

|  | Patient 1 | Patient 2 | Patient 3 | Patient 4 | Patient 5 | Patient 6 | Patient 7 | Patient 8 | Patient 9 | Patient 10 | Patient 11 | Patient 12 |
| --- | --- | --- | --- | --- | --- | --- | --- | --- | --- | --- | --- | --- |
| Race | Other | White | Other | Other | White | Other | White | Other | Other | Other | Unknown | White |
| Ethnicity | Hispanic | Non-Hispanic | Hispanic | Non-Hispanic | Non-Hispanic | Hispanic | Hispanic | Hispanic | Hispanic | Hispanic | Unknown | Non-Hispanic |
| Symptoms | Fever; Cough; Dyspnea; Nausea/Emesis; Pharyngitis; Headache | Fever; Cough; Dyspnea | Fever; Cough; Dyspnea; Nausea/Emesis; Severe Chest Pain; Abdominal Pain; Myalgias; Tachypnea | Fever; Cough; Dyspnea | Fever; Cough; Myalgias; Dyspnea | Cough; Dyspnea | Fever; Cough; Malaise; Dyspnea | Fever; Headache; Malaise; Nausea/Emesis; Diarrhea; Dyspnea; Cough | Fever; Dyspnea | Fever; Myalgias; Dyspnea | Dyspnea | Dyspnea, Fever, Cough |
| Highest Level of Oxygen Therapy | ECMO | ECMO | ECMO | Mechanical Ventilation | Mechanical Ventilation | ECMO | High Flow Nasal Cannula | ECMO | ECMO | ECMO | ECMO | ECMO |
| Number of MSC Infusions | 3 | 3 | 3 | 2 | 2 | 2 | 3 | 3 | 3 | 3 | 3 | 3 |
| COVID Diagnosis | All patients were positive for SARS-CoV-2 virus |  |  |  |  |  |  |  |  |  |  |  |
| Test results on hospital admission |  |  |  |  |  |  |  |  |  |  |  |  |
| White Blood Cell Count | 6.1 | 10.7 | 24.73 | 6.07 | 3.3 | 3.6 | 2.9 | 6.5 | 6.8 | 12.2 | NA | 9.8 |
| Neutrophil Percentage | 64.7 | NA | 85.9 | 79.7 | 80 | 52.2 | 64.5 | 76.9 | 74.1 | 78.6 | NA | NA |
| Neutrophil Count | 3.94 | NA | 21.2 | 4.84 | 2.6 | 1.19 | 1.9 | 5.43 | 3.3 | 9.56 | NA | NA |
| Lymphocyte Percentage | 27.5 | 7 | 3.5 | 15.3 | 12.6 | 28.60 | 25.2 | 13.2 | 20 | 8.6 | NA | NA |
| Lymphocyte Count | NA | 0.7 | 0.9 | 0.93 | 0.41 | 1 | 0.7 | 0.93 | 0.9 | 1.04 | NA | NA |
| Hemoglobin | 15.1 | 13.6 | 11.4 | 11.1 | 14.3 | 12 | 13 | 9.7 | 14.1 | 8.4 | NA | NA |
| Platelet Count | 239 | 184 | 602 | 278 | 167 | 136 | 155 | 278 | 269 | 503 | NA | NA |
| Prothrombin Time | NA | NA | 12.1 | NA | NA | NA | NA | NA | 13.5 | NA | NA | NA |
| Albumin | 3.3 | 2.9 | 2.7 | 3.7 | 4.8 | 4.1 | 3.8 | 3.1 | 3.9 | 3.1 | NA | NA |
| Creatinine Kinase | NA | NA | NA | 156 | NA | NA | NA | 46 | NA | 59 | NA | 1.1 |
| Alanine Aminotransferase | 110 | 30 | 58 | 20 | 18 | 22 | 31 | 19 | 68 | 21 | NA | NA |
| Aspartate Aminotransferase | 138 | 40 | 56 | 34 | 24 | 25 | 38 | 30 | 68 | 24 | NA | NA |
| Total bilirubin | 0.5 | 0.8 | 0.4 | 0.3 | 0.3 | 0.4 | 0.8 | 0.3 | 0.31 | 0.7 | NA | NA |
| Sodium | 133 | 131 | 138 | 130 | 139 | 136 | 136 | 137 | 132 | 138 | NA | 138 |
| Potassium | 3.7 | 3.7 | 3.6 | 4.4 | 3.5 | 5.2 | 4 | 3.4 | 3.6 | 3.4 | NA | 4.6 |
| Creatinine | 0.6 | 1 | 0.53 | 0.88 | 0.8 | 0.55 | 0.6 | 0.5 | 0.8 | 0.4 | NA | 0.6 |
| C-Reactive Protein | NA | NA | NA | 4.24 | 8.5 | NA | 9 | 262.5 | 17.09 | 256.7 | NA | 1.5 |
| Lactate | 2.1 | NA | 1.2 | NA | 1.3 | NA | NA | NA | NA | NA | NA | NA |

**Suplement Figure 1.**
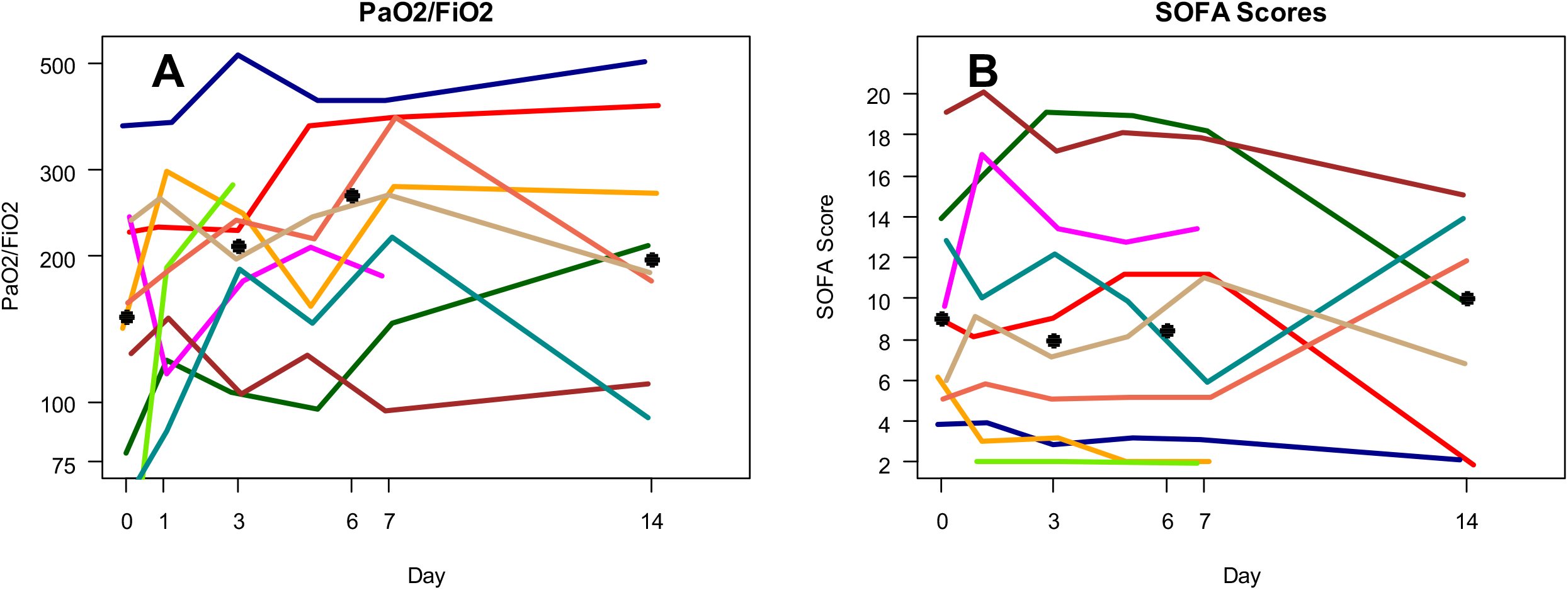
Temporal changes of PaO2/FiO2 and SOFA score in patients receiving MSC treatment. A. Change in PaO_2_/FiO_2_ of the MSC treated patients from day 0 to day 14 after treatment. B. Change in Sequential Organ Failure Assessment (SOFA) score of the patients with MSC treatment (range 0-24, with higher scores indicting more severe illness, see footnote to Table 2 for more complete definition).

**Supplement Figure 2.**
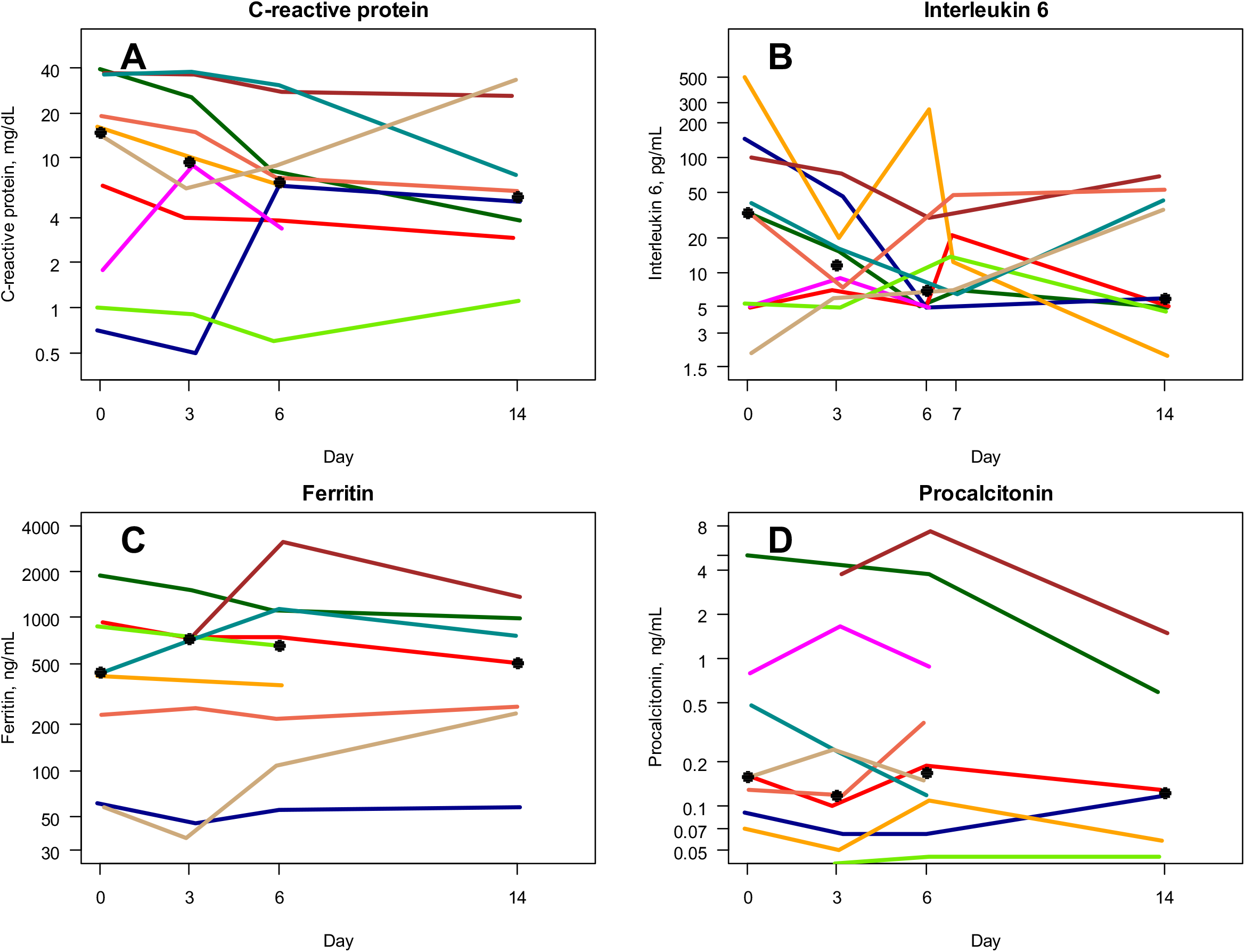
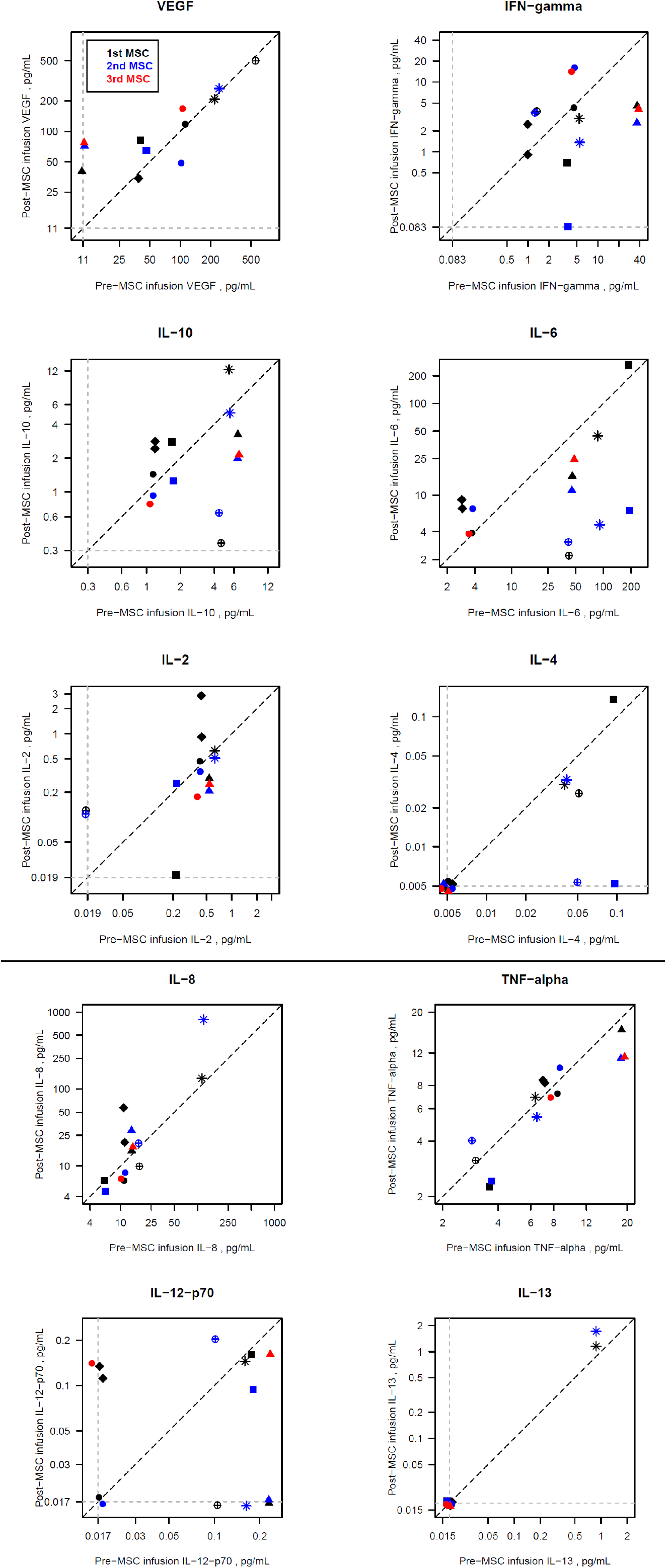
Temporal changes of (A) CRP, (B) ferritin, (C) Interleukin 6, and (D) procalcitonin in patients receiving MSC treatment.

**Supplement Figure 3.**
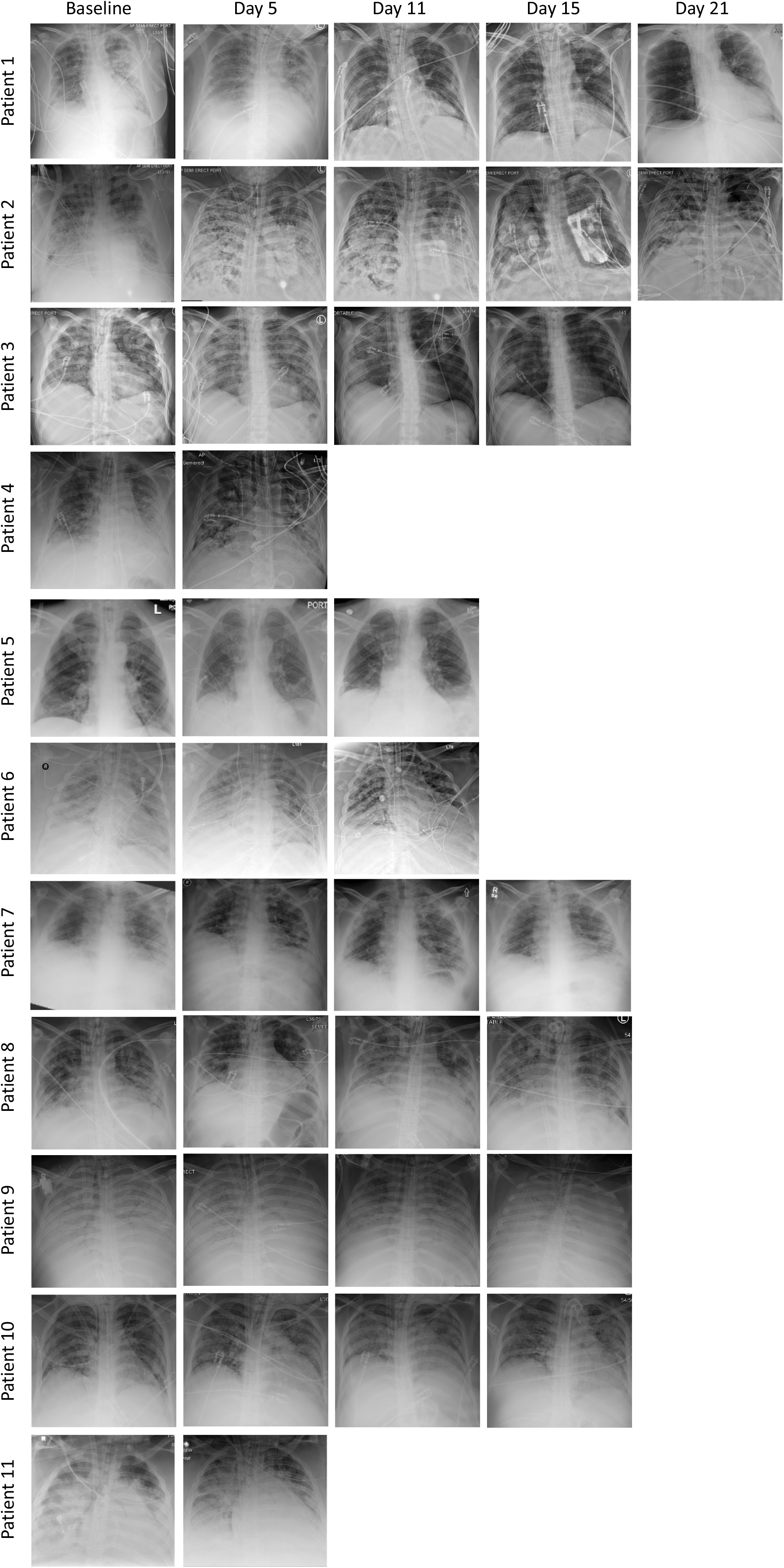
Chest radiograph images of the critically severe COVID-19 patients at baseline and interval times after MSC treatment.

## References

1. Huang C, Wang Y, Li X, Ren L, Zhao J, Hu Y, Zhang L, Fan G, Xu J, Gu X, Cheng Z, Yu T, Xia J, Wei Y, Wu W, Xie X, Yin W, Li H, Liu M, Xiao Y, Gao H, Guo L, Xie J, Wang G, Jiang R, Gao Z, Jin Q, Wang J, Cao B. Clinical features of patients infected with 2019 novel coronavirus in Wuhan, China. Lancet. 2020;395(10223):497–506. Epub 2020/01/28. doi: 10.1016/s0140-6736(20)30183-5. PubMed PMID: 31986264; PubMed Central PMCID: PMCPmc7159299.

2. Wu Z, McGoogan JM. Characteristics of and Important Lessons From the Coronavirus Disease 2019 (COVID-19) Outbreak in China: Summary of a Report of 72314 Cases From the Chinese Center for Disease Control and Prevention. Jama. 2020. Epub 2020/02/25. doi: 10.1001/jama.2020.2648. PubMed PMID: 32091533.

3. Bourgonje AR, Abdulle AE, Timens W. Angiotensin-converting enzyme-2 (ACE2), SARS-CoV-2 and pathophysiology of coronavirus disease 2019 (COVID-19)2020. doi: 10.1002/path.5471. PubMed PMID: 32418199.

4. Fehr AR, Channappanavar R, Perlman S. Middle East Respiratory Syndrome: Emergence of a Pathogenic Human Coronavirus. Annual review of medicine. 2017;68:387–99. Epub 2016/08/31. doi: 10.1146/annurev-med-051215-031152. PubMed PMID: 27576010; PubMed Central PMCID: PMCPmc5353356.

5. Chen G, Wu D, Guo W, Cao Y, Huang D, Wang H, Wang T, Zhang X, Chen H, Yu H, Zhang X, Zhang M, Wu S, Song J, Chen T, Han M, Li S, Luo X, Zhao J, Ning Q. Clinical and immunological features of severe and moderate coronavirus disease 2019. The Journal of clinical investigation. 2020;130(5):2620–9. Epub 2020/03/29. doi: 10.1172/jci137244. PubMed PMID: 32217835; PubMed Central PMCID: PMCPmc7190990.

6. Ruan Q, Yang K. Clinical predictors of mortality due to COVID-19 based on an analysis of data of 150 patients from Wuhan, China 2020;46(5):846–8. doi: 10.1007/s00134-020-05991-x. PubMed PMID: 32125452.

7. Moore JB, June CH. Cytokine release syndrome in severe COVID-19. Science. 2020;368(6490):473–4. Epub 2020/04/19. doi: 10.1126/science.abb8925. PubMed PMID: 32303591.

8. Lu H. Drug infusion options for the 2019-new coronavirus (2019-nCoV). Bioscience trends. 2020;14(1):69–71. Epub 2020/01/31. doi: 10.5582/bst.2020.01020. PubMed PMID: 31996494.

9. Grein J, Ohmagari N, Shin D, Diaz G, Asperges E, Castagna A, Feldt T, Green G, Green ML, Lescure FX, Nicastri E. Compassionate Use of Remdesivir for Patients with Severe Covid-192020. doi: 10.1056/NEJMoa2007016. PubMed PMID: 32275812.

10. Wu C, Chen X, Cai Y, Xia J, Zhou X, Xu S, Huang H, Zhang L, Zhou X, Du C, Zhang Y, Song J, Wang S, Chao Y, Yang Z, Xu J, Zhou X, Chen D, Xiong W, Xu L, Zhou F, Jiang J, Bai C, Zheng J, Song Y. Risk Factors Associated With Acute Respiratory Distress Syndrome and Death in Patients With Coronavirus Disease 2019 Pneumonia in Wuhan, China. JAMA internal medicine. 2020. Epub 2020/03/14. doi: 10.1001/jamainternmed.2020.0994. PubMed PMID: 32167524; PubMed Central PMCID: PMCPmc7070509.

11. Henry BM, Lippi G. Poor survival with extracorporeal membrane oxygenation in acute respiratory distress syndrome (ARDS) due to coronavirus disease 2019 (COVID-19): Pooled analysis of early reports. Journal of critical care. 2020;58:27–8. Epub 2020/04/13. doi: 10.1016/j.jcrc.2020.03.011. PubMed PMID: 32279018; PubMed Central PMCID: PMCPmc7118619.

12. Volarevic V, Gazdic M, Simovic Markovic B, Jovicic N, Djonov V, Arsenijevic N. Mesenchymal stem cell-derived factors: Immuno-modulatory effects and therapeutic potential. Biofactors. 2017;43(5):633–44. Epub 2017/07/19. doi: 10.1002/biof.1374. PubMed PMID: 28718997.

13. Pittenger MF, Discher DE, Péault BM, Phinney DG. Mesenchymal stem cell perspective: cell biology to clinical progress2019;4:22. doi: 10.1038/s41536-019-0083-6. PubMed PMID: 31815001.

14. Hare JM, Fishman JE, Gerstenblith G, DiFede Velazquez DL, Zambrano JP, Suncion VY, Tracy M, Ghersin E, Johnston PV, Brinker JA, Breton E, Davis-Sproul J, Schulman IH, Byrnes J, Mendizabal AM, Lowery MH, Rouy D, Altman P, Wong Po Foo C, Ruiz P, Amador A, Da Silva J, McNiece IK, Heldman AW, George R, Lardo A. Comparison of allogeneic vs autologous bone marrow–derived mesenchymal stem cells delivered by transendocardial injection in patients with ischemic cardiomyopathy: the POSEIDON randomized trial. Jama. 2012;308(22):2369–79. Epub 2012/11/03. doi: 10.1001/jama.2012.25321. PubMed PMID: 23117550; PubMed Central PMCID: PMCPmc4762261.

15. Leng Z, Zhu R, Hou W, Feng Y, Yang Y, Han Q, Shan G, Meng F, Du D, Wang S, Fan J, Wang W, Deng L, Shi H, Li H, Hu Z, Zhang F, Gao J, Liu H, Li X, Zhao Y, Yin K, He X, Gao Z, Wang Y, Yang B, Jin R, Stambler I, Lim LW, Su H, Moskalev A, Cano A, Chakrabarti S, Min KJ, Ellison-Hughes G, Caruso C, Jin K, Zhao RC. Transplantation of ACE2(-) Mesenchymal Stem Cells Improves the Outcome of Patients with COVID-19 Pneumonia. Aging and disease. 2020;11(2):216–28. Epub 2020/04/08. doi: 10.14336/ad.2020.0228. PubMed PMID: 32257537; PubMed Central PMCID: PMCPmc7069465.

16. Alshahrani MS, Sindi A, Alshamsi F, Al-Omari A, El Tahan M, Alahmadi B, Zein A, Khatani N, Al-Hameed F, Alamri S, Abdelzaher M, Alghamdi A, Alfousan F, Tash A, Tashkandi W, Alraddadi R, Lewis K, Badawee M, Arabi YM, Fan E, Alhazzani W. Extracorporeal membrane oxygenation for severe Middle East respiratory syndrome coronavirus. Annals of intensive care. 2018;8(1):3. Epub 2018/01/14. doi: 10.1186/s13613-017-0350-x. PubMed PMID: 29330690; PubMed Central PMCID: PMCPmc5768582.

17. Chen J, Hu C, Chen L, Tang L, Zhu Y, Xu X, Chen L, Gao H, Lu X, Yu L, Dai X, Xiang C, Li L. Clinical study of mesenchymal stem cell treating acute respiratory distress syndrome induced by epidemic Influenza A (H7N9) infection, a hint for COVID-19 infusion. Engineering (Beijing, China). 2020. Epub 2020/04/16. doi: 10.1016/j.eng.2020.02.006. PubMed PMID: 32292627; PubMed Central PMCID: PMCPmc7102606.

